# Cultryx: Precision Diagnostic Stewardship for Blood Cultures Using Machine Learning

**DOI:** 10.64898/2026.02.27.26347214

**Authors:** Nicholas P. Marshall, Wenyuan Chen, Fatemeh Amrollahi, Fateme Nateghi Haredasht, Manoj V. Maddali, Stephen P. Ma, Aydin Zahedivash, Kameron C. Black, Amy Chang, Stanley C. Deresinski, Mary K. Goldstein, Steven M Asch, Niaz Banaei, Jonathan H Chen

**Affiliations:** Division of Pediatric Infectious Diseases, Department of Pediatrics, School of Medicine, Stanford University, Palo Alto, California, USA; Division of Computational Medicine, Stanford University, Stanford, California, USA; Division of Pulmonary and Critical Care, Department of Medicine, School of Medicine, Stanford University, Stanford, California, USA; Division of Hospital Medicine, Department of Medicine, School of Medicine, Stanford University, Stanford, California, USA; Division of Clinical Informatics, Department of Pediatrics, School of Medicine, Stanford University, Palo Alto, California, USA; Division of Infectious Diseases and Geographic Medicine, Department of Medicine, School of Medicine, Stanford University, Stanford, California, USA; Department of Health Policy, School of Medicine, Stanford University, Stanford, California, USA; Division of Primary Care and Population Health, Department of Medicine, School of Medicine, Stanford University, Stanford, California, USA; Department of Pathology, School of Medicine, Stanford University, Stanford, California, USA; Clinical Excellence Research Center, School of Medicine, Stanford University, Palo Alto, California, USA

**Keywords:** blood culture stewardship, diagnostic stewardship, bacteremia prediction, machine learning, large language model, clinical decision support, electronic health record, clinical informatics, resource conservation

## Abstract

**Background:** The 2024 blood culture bottle shortage brought diagnostic resource allocation to the forefront, reflecting persistent, foundational challenges with low-value testing and empiric treatment approaches under clinical uncertainty.

**Objective:** To determine whether a machine learning approach using electronic medical record data can predict bacteremia more effectively than existing systems and practices to guide diagnostic testing and empiric treatment strategies.

**Methods:** In a retrospective cohort of 101,812 adult emergency department encounters (2015-2025), we first established an idealized cognitive baseline by evaluating physician and generative AI (GPT-5) application of the professional society-endorsed Fabre framework on a validation subset. We then trained an XGBoost model (Cultryx) on the full cohort to predict bacteremia, benchmarking its performance against real-world clinical heuristics (SIRS, Shapiro Rule).

**Results:** For the idealized baseline, physicians applying the Fabre framework achieved 95.7% sensitivity, but GPT-5 automation failed to replicate this standard (71.6% sensitivity). In real-world benchmarking, Cultryx outperformed all clinical heuristics (AUROC 0.810). SIRS lacked specificity (41.2%), driving diagnostic overuse, while the Shapiro Rule lacked sensitivity (70.2%), missing ~30% of bacteremia cases. In contrast, when calibrated to a strict 95% sensitivity target, Cultryx achieved the highest culture volume deferral rate (26.2%, deferring ~ 15,872 bottles with predicted negative results) while maintaining a 98.9% negative predictive value. Cultryx^score^, a simplified bedside tool, retained a 20.8% deferral rate.

**Conclusions:** Machine learning provides a superior, data-driven alternative to mainstream clinical heuristics for predicting bacteremia. By maximizing culture deferment without compromising pathogen detection, Cultryx can conserve diagnostic resources, reduce unnecessary empiric antibiotic exposure, and systematically elevate patient safety.

**Summary:** Cultryx, a machine learning model for blood culture stewardship, outperforms standard clinical heuristics in predicting bacteremia. This approach could reduce culture utilization by over 26% while preserving pathogen detection, conserving diagnostic resources, reducing unnecessary antibiotic exposure, and elevating patient safety.

## 1 Introduction

Timely diagnosis of bacteremia is a critical task in acute care, as delays can lead to rapid clinical deterioration and death. Blood cultures remain the diagnostic gold standard, uniquely providing organism identification and antimicrobial susceptibility data to guide both agent selection and treatment duration.[1] Their diagnostic yield, however, declines substantially after antibiotic administration, mandating collection prior to treatment.[2] Despite their importance, blood cultures are frequently overused; with true pathogen positivity rates below 10% and as many as 60% obtained without strong clinical indication, this low-threshold practice initiates a cascade of unnecessary hospitalizations, excess antimicrobial exposure, and iatrogenic harm, while placing significant strain on clinical and laboratory resources.[1, 3] This overuse is driven by the diagnostic uncertainty clinicians face when initiating empiric therapy while awaiting culture results, a process that can take up to five days.[1]

The inherent vulnerability of this diagnostic strategy was exposed during the 2024 global shortage of BD BACTEC™ blood culture bottles.[4, 5] What began as a supply chain disruption evolved into a diagnostic crisis, affecting nearly 76% of facilities using the system.[6] Hospitals were forced to implement crude rationing strategies, including hard stops and single-set collections, resulting in a 27.4% precipitous decline in culture volume at affected centers.[5, 6, 7] Most alarmingly, this indiscriminate reduction in testing led to a 15.3% decrease in the detection of confirmed BSIs, highlighting the severe patient safety risks inherent in non-targeted rationing.[6]

Recent work has explored AI-driven decision support to improve diagnostic stewardship, using EHR-based machine-learning risk models and, more recently, large language models (LLMs) to synthesize pre-test clinical context.[8, 9, 10, 11] However, reproducibility and real-world utility of these approaches for guiding blood culture ordering, especially under resource constraints, remain insufficiently validated.

This crisis highlighted a critical gap in clinical practice — the absence of scalable, data-driven tools to guide pre-test probability assessment. While alternative diagnostic modalities such as rapid molecular assays have emerged, they remain adjuncts that cannot replace the susceptibility data provided by culture required for definitive therapy.[12, 13] Consequently, the most effective strategy for resilience is not to replace the blood culture, but to refine the decision to obtain one.[5, 14, 15] To date, this refinement has relied on expert consensus recommendations, most notably the Fabre framework.[3] Derived from a comprehensive scoping review of 50 studies, this framework categorizes clinical syndromes into low (*<*10%), moderate (10–50%), and high (*>*50%) pre-test probabilities of bacteremia.[3] During the supply chain crisis, the Fabre framework was one of the prominent strategies recommended by the Infectious Diseases Society of America (IDSA), the Society for Healthcare Epidemiology of America (SHEA), and the American Society for Microbiology (ASM) for rationing, guiding clinicians to prioritize testing for high-risk phenotypes while deferring cultures for low-risk presentations.[4, 5]

In this study, we first assessed the scalability of this expert-driven approach. To establish an idealized cognitive baseline, we evaluated the performance of human physicians applying the Fabre framework retrospectively, with full access to clinical data. We then tested whether this expert performance could be replicated at scale using generative AI (GPT-5). Recognizing that manual application of complex frameworks is fundamentally impractical in the fast-paced workflow of a busy emergency department (ED) and anticipating the safety limitations of uncalibrated LLMs, we sought to develop a purpose-built, automated solution. We leveraged the Stanford Antibiotic Resistance Microbiology Dataset (ARMD) to train and validate Cultryx, a machine learning model (XGBoost) designed to seamlessly predict bacteremia at the time of order using structured electronic health record data.[16, 17] To ensure these insights translate to resource-limited settings or system downtimes, we also derived the Cultryx^score^, a simplified integer-based bedside calculator distilled directly from the parent model’s strongest predictors.

Real-world ED triage is characterized by inherent time constraints and frequently incomplete clinical information, rendering the application of an idealized, comprehensive framework impractical for frontline providers. Consequently, rather than benchmarking Cultryx against the Fabre framework, we evaluated its performance against the standard clinical decision rules clinicians rely on in practice. Specifically, we compared its performance against two established tools: the Systemic Inflammatory Response Syndrome (SIRS) criteria and the Shapiro Rule.[18, 19, 20, 21] While SIRS is highly sensitive for detecting physiological stress, it lacks the specificity required to distinguish bacteremia from non-infectious inflammation.[18] Conversely, the Shapiro Rule is tailored to identify low-risk ED patients who may safely forgo culture collection.[19, 20, 21] By comparing Cultryx and Cultryx^score^ against these practical baselines, we aimed to assess whether precision machine learning offers a more effective strategy than traditional decision aids for guiding safe, sustainable blood culture stewardship.

## 2 Methods

### 2.1 Study Design and Data Source

We conducted a retrospective cohort study using deidentified electronic health record (EHR) data from the Stanford Medicine Research Data Repository (STARR), which captures longitudinal data from Stanford Health Care and affiliated hospitals.[16, 17, 22] The study protocol was approved by the Stanford University Institutional Review Board (IRB #70466) with a waiver of informed consent.

### 2.2 Cohort Generation

We identified all blood culture orders placed in the ED for adult patients (≥18 years) between 2015 and 2025. To ensure the independence of clinical events and avoid confounding by recent infection, we excluded encounters if the patient had a positive blood culture in the preceding 14 days. The final analytic cohort was split temporally based on the index order year: Training (2015-2022), Validation (2023), and Test (2024-2025).

### 2.3 Outcome Definition

The primary outcome was encounter-level bacteremia, defined as any positive blood culture set obtained during an ED encounter. A blood culture set typically consists of two bottles, drawn simultaneously. We developed a hierarchical classification algorithm to label sets as positive, contaminated, or negative. Sets were classified as positive if they demonstrated growth of a non-contaminant pathogen (e.g., *E. coli, S. aureus*). Consistent with Stanford Health Care Laboratory Policy, we defined a contaminant as a single blood culture set (representing a single venipuncture) growing skin flora, specifically: coagulase-negative Staphylococcus, viridans group streptococci, and/or Gram-positive rods (e.g., *Corynebacterium* spp., *Bacillus* spp., *Cutibacterium acnes*).[23] To avoid misclassifying true pathogens, we applied a promotion rule: potentially contaminating organisms isolated in ≥2 sets within the same encounter (without explicit contamination comments) were promoted to positive. Sets with no growth or explicit negative test results were labeled negative. An encounter was labeled positive if at least one set met the positivity criteria; encounters with only contaminant growth were treated as negative/non-bacteremic for the primary prediction analysis.

### 2.4 Clinical Features and Preprocessing

We extracted demographic data, vital signs, and laboratory results available in the 48 hours preceding the index blood culture. Features were harmonized and aggregated (minimum, maximum, median) to the encounter level. The final model utilized 36 clinical predictors, including demographics (age, sex, BMI, invasive devices), vital sign extremes (temperature, heart rate, blood pressure, respiratory rate, oxygen saturation), and comprehensive laboratory markers (complete blood count, metabolic panels, lactate, and C-reactive protein). We additionally derived composite features, such as the shock index and binary indicators for hemodynamic instability (e.g., hypotension or hyperlactatemia). Missing data were handled natively by the tree-based model; for linear baselines, we utilized median imputation derived from the training set. A complete list of features are provided in the Supplementary Appendix A.

### 2.5 Experiment 1: Establishing an Idealized Cognitive Baseline with Human and AI Application of the Fabre Framework

To evaluate the scalability of expert-driven stewardship, we assessed the reproducibility of the Fabre framework when applied by a large language model (LLM) versus human experts retrospectively. We selected a stratified random sample of 112 encounters from the test set.[3] To ensure robust evaluation of model sensitivity given the low baseline prevalence of bacteremia (*<*10%), we oversampled culture-positive cases to comprise 80% of this validation subset. These encounters underwent independent manual chart review (simulating an idealized environment with full data availability) by two board-certified physicians to establish a “ground truth” risk tier (low, intermediate, or high). Disagreements were adjudicated by an infectious diseases specialist to generate a final consensus label. We then provided a HIPAA-compliant GPT-5 model with the identical pre-test clinical context (via a structured prompt) and instructed it to assign risk tiers; the full prompt text is available in Supplementary Appendix B. We assessed the concordance between the LLM and the physician consensus by calculating Cohen’s *κ*, as well as the sensitivity and specificity of the LLM for identifying encounters where the expert framework recommended blood cultures (intermediate or high risk) relative to the clinician reference standard.

### 2.6 Experiment 2: Benchmarking Against Real-World Operational Heuristics

Because comprehensive frameworks are often impractical in the time-constrained ED environment, we benchmarked our model against established clinical standards that clinicians actively rely on in practice. We evaluated the performance of the SIRS criteria and the Shapiro Rule as binary predictors of bacteremia.[18, 19] For the Shapiro Rule, which originally includes subjective clinical assessments (e.g., “suspicion of endocarditis”), we implemented a modified version utilizing only objective, numerical components available in structured EHR data (e.g., temperature, neutrophil count, platelet count) to ensure scalable, automated calculation. For both rules, scores were computed using the most abnormal values recorded within the 48-hour window preceding the index blood culture order.

### 2.7 Experiment 3: Machine Learning and Simplified Bedside Tool

We trained a Gradient Boosted Tree (XGBoost) model, termed Cultryx, to predict encounter-level bacteremia using 36 structured clinical features. To optimize performance while preventing overfitting, hyperparameters, including learning rate, maximum tree depth, and regularization terms, were tuned via randomized search with 5-fold cross-validation on the training set. Given the significant class imbalance (7.5% culture positivity rate), we applied positive class weighting inversely proportional to the class frequency during the training objective function to ensure adequate sensitivity for the minority class.

To facilitate implementation in settings lacking an integrated ML infrastructure, we derived a transparent, integer-based risk score, termed Cultryx^score^. We computed SHAP (SHapley Additive exPlanations) values for the Cultryx model to identify the top 15 predictors. These continuous features were discretized into binary risk flags based on established clinical thresholds (e.g., Temperature >38^°^C, White Blood Cell count >12×10^9^/L, Platelets <150×10^9^/L) or data-driven quartiles where standard cutoffs did not exist. We then fit a logistic regression model using these binary features; the resulting coefficients were scaled and rounded to the nearest integer to produce a summable risk score (ranging from 0 to 4 points per item).

To ensure that model outputs could safely guide stewardship, we calibrated the predicted probabilities of both Cultryx and Cultryx^score^ using Platt scaling.[24] We then determined decision thresholds on the validation set necessary to achieve pre-specified sensitivity targets (85%, 90%, 95%, and 98%), prioritizing patient safety over the reduction in testing volume. Encounters with calibrated risks below these thresholds were classified as “low risk” and deemed eligible to defer culture and potentially antibiotic orders. We quantified the clinical impact using two metrics: the “culture deferral rate” (defined as the proportion of all encounters classified as low risk and therefore eligible for testing deferral) and the estimated “bottle savings.” Because standard institutional practice requires collecting two blood culture sets (comprising four total bottles) per order, bottle savings were calculated by multiplying the absolute number of deferred encounters by four.

## 3 Results

### 3.1 Cohort

The final analytic cohort spanned 101,812 ED encounters (62,919 unique patients) from 2015 to 2025 (Table 1). The median age was 65 years (IQR 49.0-78.0), and 49.4% of encounters were among female patients.

**Table 1:** Baseline characteristics of adult emergency department encounters undergoing blood culture testing.

|  | All | Train | Validation | Test | Human Review Test |
| --- | --- | --- | --- | --- | --- |
| Encounters, n | 101,812 | 73,256 | 13,412 | 15,144 | 112 |
| Bacteremia, n (%) | 7,635 (7.5) | 5,682 (7.8) | 890 (6.6) | 1,063 (7.0) | 102 (91.1) |
| Contamination, n (%) | 610 (0.6) | 560 (0.8) | 25 (0.2) | 25 (0.2) | 0 (0.0) |
| Negative, n (%) | 93,567 (91.9) | 67,014 (91.5) | 12,497 (93.2) | 14,056 (92.8) | 10 (8.9) |
| Age, median (IQR), y | 65.0 (49.0–78.0) | 65.0 (49.0–78.0) | 66.0 (49.0–78.0) | 66.0 (49.0–78.0) | 69.5 (59.0–82.0) |
| Female, n (%) | 50,255 (49.4) | 36,366 (49.6) | 6,565 (48.9) | 7,324 (48.4) | 49 (43.8) |
| BMI, median (IQR), kg/m<sup>2</sup> | 25.3 (21.8–29.9) | 25.3 (21.7–29.9) | 25.4 (21.9–29.9) | 25.4 (21.9–29.8) | 25.0 (21.2–29.4) |
| Any invasive line, n (%) | 4,856 (4.8) | 3,838 (5.2) | 497 (3.7) | 521 (3.4) | 9 (8.0) |
| Temp max, median (IQR), °C | 37.0 (36.7–37.6) | 37.0 (36.7–37.6) | 36.9 (36.6–37.4) | 36.9 (36.6–37.5) | 37.3 (36.7–38.1) |
| HR max, median (IQR), beats/min | 99 (86–115) | 99 (85–115) | 99 (85–115) | 101 (86–116) | 110.0 (94.0–124.0) |
| SBP min, median (IQR), mmHg | 117 (103–133) | 117 (103–133) | 117 (103–133) | 115 (101–131) | 107.5 (93.2–125.8) |
| Resp max, median (IQR), breaths/min | 20 (18–24) | 20 (18–24) | 20 (18–24) | 20 (18–25) | 21.0 (18.0–26.0) |
| SpO<sub>2</sub> min, median (IQR), % | 96 (94–98) | 96 (94–98) | 96 (94–98) | 96 (93–98) | 96.0 (93.0–98.0) |
| Shock index, median (IQR) | 0.8 (0.7–1.0) | 0.8 (0.7–1.0) | 0.8 (0.7–1.0) | 0.9 (0.7–1.1) | 1.0 (0.8–1.3) |
| WBC max, median (IQR), 10<sup>3</sup>/μL | 10.0 (7.0–14.3) | 10.1 (7.0–14.4) | 9.8 (7.0–14.0) | 10.1 (7.0–14.4) | 11.3 (8.1–17.4) |
| ANC max, median (IQR), 10<sup>3</sup>/μL | 7.6 (4.9–11.4) | 7.6 (4.9–11.5) | 7.4 (4.9–11.0) | 7.7 (5.0–11.5) | 10.2 (6.7–14.5) |
| Lymphocytes min, median (IQR), % | 11.0 (6.1–18.7) | 10.9 (6.0–18.5) | 11.6 (6.5–19.1) | 11.2 (6.1–18.9) | 5.1 (3.2–8.1) |
| Hemoglobin min, median (IQR), g/dL | 11.5 (9.7–13.1) | 11.5 (9.7–13.1) | 11.6 (9.9–13.2) | 11.7 (9.8–13.3) | 10.6 (9.1–12.3) |
| Platelets min, median (IQR), 10<sup>3</sup>/μL | 219 (158–295) | 214 (154–288) | 237 (172–315) | 230 (167–305) | 186.0 (129.0–306.0) |
| Lactate max, median (IQR), mmol/L | 1.5 (1.0–2.3) | 1.5 (1.0–2.3) | 1.5 (1.0–2.3) | 1.5 (1.0–2.3) | 2.0 (1.5–3.1) |
| Creatinine max, median (IQR), mg/dL | 1.0 (0.7–1.4) | 1.0 (0.8–1.4) | 0.9 (0.7–1.3) | 0.9 (0.7–1.3) | 1.1 (0.8–1.6) |
| CRP max, median (IQR), mg/dL | 5.6 (1.8–12.9) | 5.8 (1.8–13.5) | 4.9 (1.6–11.3) | 5.1 (1.8–11.8) | 19.3 (10.5–21.1) |

Abbreviations: BMI, body mass index; WBC, white blood cell count; ANC, absolute neutrophil count; CRP, C-reactive protein; IQR, interquartile range.

The overall prevalence of bacteremia at the encounter level was 7.5%, while 0.6% of encounters represented contamination, and 91.9% were negative. Among positive cultures, *Escherichia coli* was the most frequently isolated pathogen, followed by *Staphylococcus aureus* and *Klebsiella pneumoniae*. Contaminant cultures were largely comprised of coagulase-negative Staphylococcus species (Figure 1).

**Figure 1.**
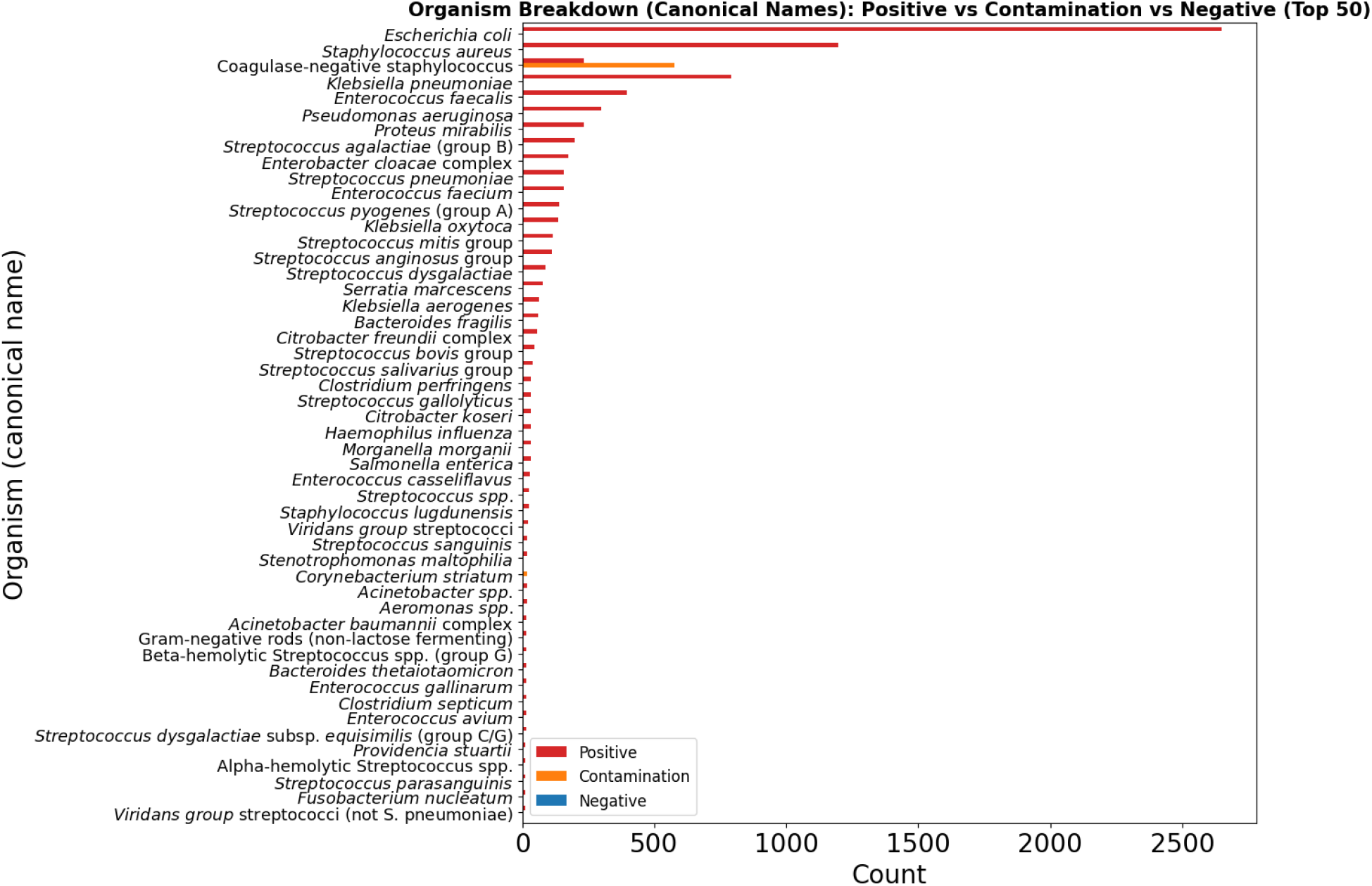
Organism Breakdown (Canonical Names): Positive vs Contamination vs Negative (Top 50)

### 3.2 Experiment 1: Establishing an Idealized Cognitive Baseline with Human and AI Application of the Fabre Framework

In the subset of 112 encounters undergoing manual chart review, the Fabre framework demonstrated substantial interrater reliability between human experts, yielding a Cohen’s *κ* of 0.733 (95% CI, 0.598-0.853). However, when the HIPAA-compliant GPT-5 model was prompted with the same clinical data, it achieved only fair agreement with the physician consensus (*κ* = 0.363; 95% CI, 0.165-0.555).

Regarding performance metrics relative to the adjudicated consensus (Table 2), human reviewers maintained a sensitivity of 95.7% for identifying high-risk encounters. In contrast, the LLM achieved a sensitivity of 71.6% and a Negative Predictive Value (NPV) of 0.121 relative to the expert standard. Specificity was 35.0% for human reviewers and 40.0% for the LLM.

**Table 2:**
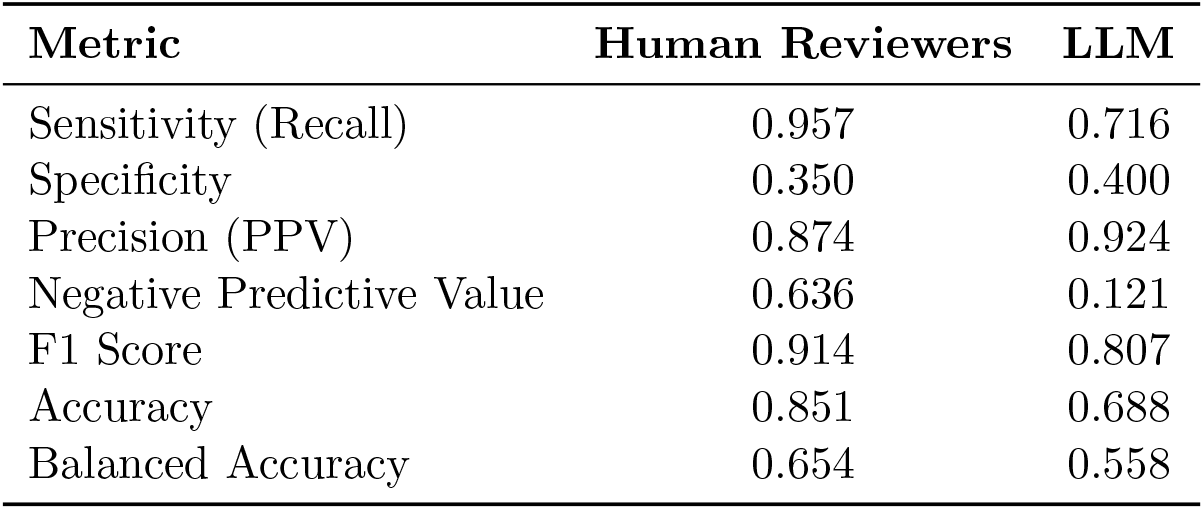
Comparison of Human and LLM Performance for Identifying Encounters Requiring Blood Cultures (Fabre Intermediate or High Risk)

| Metric | Human Reviewers | LLM |
| --- | --- | --- |
| Sensitivity (Recall) | 0.957 | 0.716 |
| Specificity | 0.350 | 0.400 |
| Precision (PPV) | 0.874 | 0.924 |
| Negative Predictive Value | 0.636 | 0.121 |
| F1 Score | 0.914 | 0.807 |
| Accuracy | 0.851 | 0.688 |
| Balanced Accuracy | 0.654 | 0.558 |

### 3.3 Experiment 2: Benchmarking Against Real-World Operational Heuristics

Performance metrics for established clinical rules are detailed in Table 3. The standard SIRS criteria (≥2) achieved a sensitivity of 80.2% and specificity of 41.2% (PPV 9.3%). Expanding the definition to include hemodynamic instability (hypotension or hyperlactatemia) increased sensitivity to 86.3% and decreased specificity to 33.4%.

**Table 3:** Performance of rule-based clinical baselines for predicting bacteremia in the held-out test set (15,144 emergency department encounters; prevalence 7.0%).

| Rule | TN | FP | FN | TP | Sensitivity, % | Specificity, % | PPV, % | NPV, % |
| --- | --- | --- | --- | --- | --- | --- | --- | --- |
| SIRS $\geq 2$ criteria | 5,796 | 8,285 | 210 | 853 | 80.2 | 41.2 | 9.3 | 96.5 |
| SIRS $\geq 2$ or low BP / high lactate | 4,702 | 9,379 | 146 | 917 | 86.3 | 33.4 | 8.9 | 97.0 |
| SIRS $\geq 1$ criterion | 2,043 | 12,038 | 33 | 1,030 | 96.9 | 14.5 | 7.9 | 98.4 |
| Any SIRS or low BP / high lactate | 1,772 | 12,309 | 26 | 1,037 | 97.5 | 12.6 | 7.8 | 98.6 |
| Shapiro $\geq 2$ criteria | 8,191 | 5,890 | 317 | 746 | 70.2 | 58.2 | 11.2 | 96.3 |
| Shapiro $\geq 1$ criterion | 3,265 | 10,816 | 67 | 996 | 93.7 | 23.2 | 8.4 | 98.0 |

Abbreviations: TN, true negative; FP, false positive; FN, false negative; TP, true positive; PPV, positive predictive value; NPV, negative predictive value; BP, blood pressure; SIRS, systemic inflammatory response syndrome.

The Shapiro Rule (≥2 criteria) yielded a specificity of 58.2% and a sensitivity of 70.2%. When the threshold was lowered to ≥1 criterion, specificity decreased to 23.2% while sensitivity increased to 86.5%.

### 3.4 Experiment 3: Cultryx Performance and Stewardship Impact

Cultryx achieved an Area Under the Receiver Operating Characteristic curve (AUROC) of 0.810 and an Area Under the Precision-Recall Curve (AUPRC) of 0.307 on the held-out test set. Calibration via Platt scaling reduced the Brier score from 0.121 to 0.053 (Figure 2).

**Figure 2.**
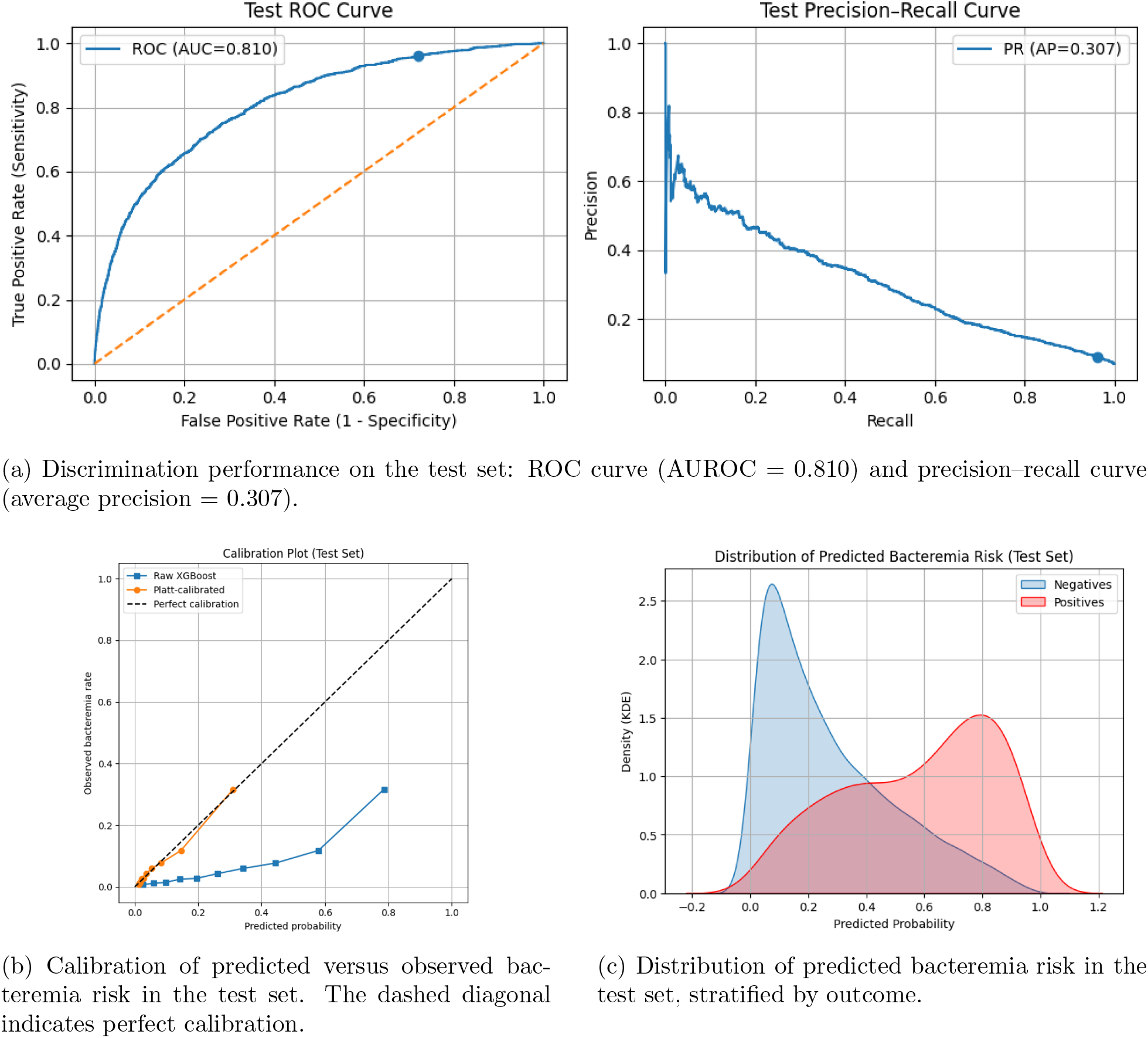
Performance of the Cultryx model on the held-out test set. (A) ROC and precision– recall curves. (B) Calibration plot comparing predicted probabilities with observed event rates. (C) Distribution of predicted risk stratified by bacteremia outcome.

We applied decision thresholds calibrated to specific sensitivity targets to estimate stewardship impact (Table 4). At a sensitivity threshold of 95%, Cultryx achieved a specificity of 27.9% and a Negative Predictive Value (NPV) of 98.9%. Applying this threshold to the test cohort resulted in a deferral rate of 26.2%, corresponding to an estimated reduction of 15,872 blood culture bottles. At a sensitivity target of 98%, Cultryx maintained a deferral rate of 12.6% (7,632 bottles saved).

**Table 4:** Comparison of calibrated Cultryx and Cultryx^score^ models across sensitivity targets on the test set. Bottles ordered and saved assume two culture sets per encounter and two bottles per set (60,576 total bottles across 15,144 test encounters; 1,066 bacteremia encounters).

| Target Sens. | Model | Sens. | Spec. | Defer Rate | Bottles Ordered | Bottles Saved | Positives Captured | Missed Positives |
| --- | --- | --- | --- | --- | --- | --- | --- | --- |
| 85% | Cultryx | 0.879 | 0.524 | 0.496 | 30,520 | 30,056 (49.6%) | 937 | 129 (12.1%) |
|  | Cultryx<sup>score</sup> | 0.872 | 0.460 | 0.437 | 34,128 | 26,448 (43.7%) | 930 | 136 (12.8%) |
| 90% | Cultryx | 0.913 | 0.434 | 0.409 | 35,776 | 24,800 (40.9%) | 973 | 93 (8.7%) |
|  | Cultryx<sup>score</sup> | 0.958 | 0.289 | 0.271 | 44,144 | 16,432 (27.1%) | 1,021 | 45 (4.2%) |
| 95% | Cultryx | 0.960 | 0.279 | 0.262 | 44,704 | 15,872 (26.2%) | 1,024 | 42 (4.0%) |
|  | Cultryx<sup>score</sup> | 0.976 | 0.222 | 0.208 | 47,984 | 12,592 (20.8%) | 1,040 | 26 (2.4%) |
| 98% | Cultryx | 0.987 | 0.135 | 0.126 | 52,920 | 7,656 (12.6%) | 1,052 | 14 (1.3%) |
|  | Cultryx<sup>score</sup> | 0.984 | 0.173 | 0.162 | 50,740 | 9,836 (16.2%) | 1,049 | 17 (1.6%) |

### 3.5 Cultryx^**score**^

Cultryx^score^, a simplified integer-based risk tool derived from SHAP values (Figure 3), identified hyperthermia (≥38^°^C), neutrophilia, thrombocytopenia, and elevated CRP as the strongest predictors of bacteremia (Table 5). This score achieved an AUROC of 0.760. When calibrated to match Cultryx’s 95% sensitivity target, Cultryx^score^ achieved a specificity of 22.2% and a deferral rate of 20.8%. This corresponds to an estimated 12,592 bottles saved, approximately 3,200 fewer than the full Cultryx model at the equivalent sensitivity threshold (Table 6).

**Table 5:**
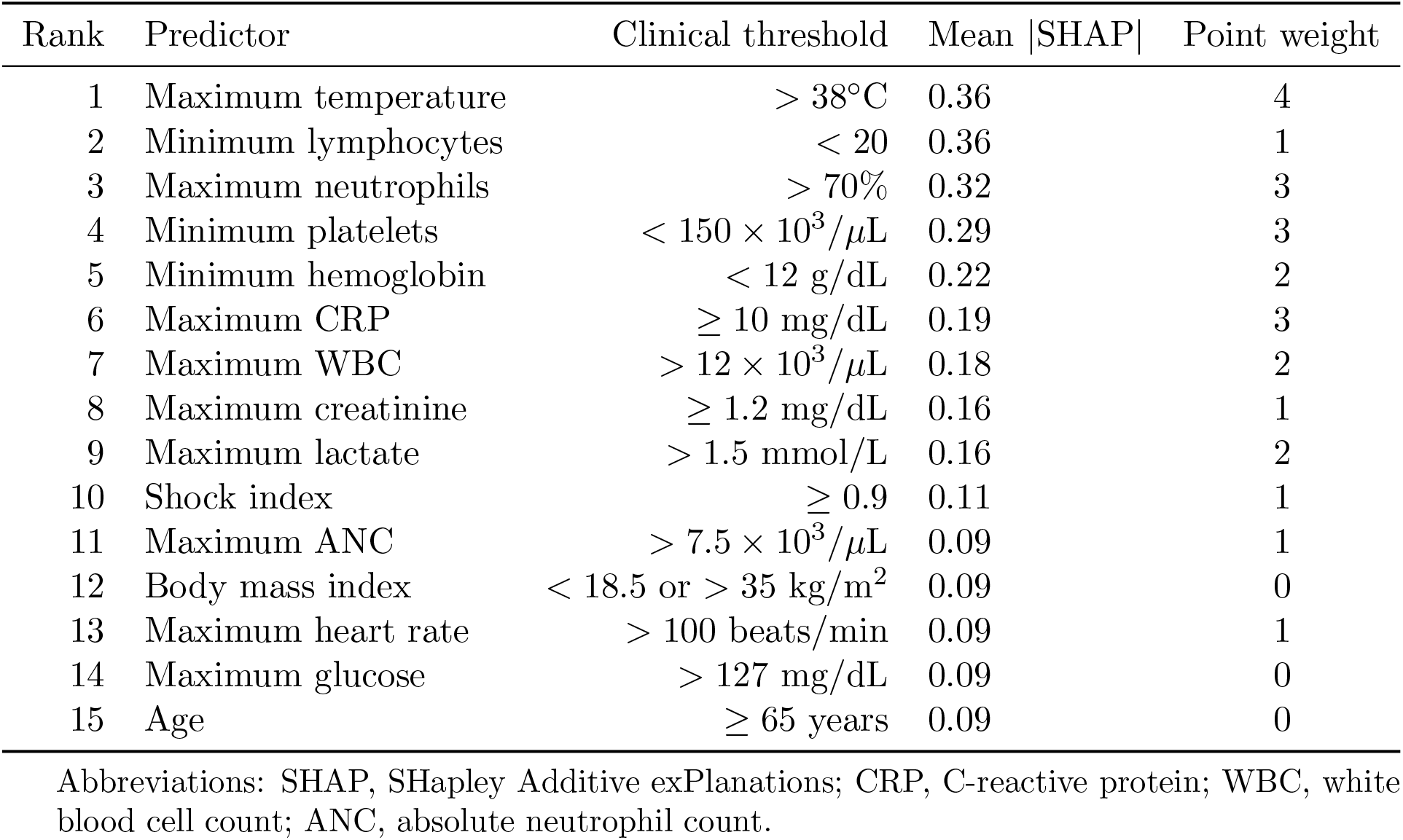
Top SHAP-ranked predictors from the Cultryx model, corresponding clinical thresholds, and point weights in the simplified bacteremia score.

**Table 6:** Performance metrics for the Cultryx, Cultryx^score^ and rule-based baselines on the held-out test set (prevalence 7.0%).

| Scenario / Model | Threshold / Cutoff | TN | FP | FN | TP | Sensitivity, % | Specificity, % | PPV, % | NPV, % | AUROC |
| --- | --- | --- | --- | --- | --- | --- | --- | --- | --- | --- |
| *Cultryx* |  |  |  |  |  |  |  |  |  |  |
| 95% sensitivity operating point | Prob. $\geq 0.102$ | 3,926 | 10,155 | 42 | 1,021 | 96.0 | 27.9 | 9.1 | 98.9 | 0.810 |
| *Cultryx<sup>score</sup>* |  |  |  |  |  |  |  |  |  |  |
| Target $\approx 95\%$ sensitivity | Points $K \geq 6$ | 3,126 | 10,955 | 26 | 1,037 | 97.6 | 22.2 | 8.6 | 99.1 | 0.760<sup>a</sup> |
| *Rule-Based Clinical Baselines* |  |  |  |  |  |  |  |  |  |  |
| SIRS $\geq 2$ criteria | N/A | 5,796 | 8,285 | 210 | 853 | 80.2 | 41.2 | 9.3 | 96.5 | N/A |
| SIRS $\geq 1$ criterion | N/A | 2,043 | 12,038 | 33 | 1,030 | 96.9 | 14.5 | 7.9 | 98.4 | N/A |
| Shapiro $\geq 1$ criterion | N/A | 3,265 | 10,816 | 67 | 996 | 93.7 | 23.2 | 8.4 | 98.0 | N/A |

Abbreviations: TN, true negative; FP, false positive; FN, false negative; TP, true positive; PPV, positive predictive value; NPV, negative predictive value; AUROC, area under the receiver operating characteristic curve.
<sup>a</sup> AUROC for the Cultryx<sup>score</sup> corresponds to discrimination of the underlying logistic model on total point scores (val AUROC 0.763; test AUROC 0.760).

**Table 7:** Final Feature Set and Definitions.

| Feature | Category | Definition |
| --- | --- | --- |
| age | Demographics/Context | Patient age at encounter (years). |
| gender_binary | Demographics/Context | Binary sex indicator (as recorded in EHR). |
| bmi | Demographics/Context | Body mass index ($\text{kg}/\text{m}^2$). |
| has_any_line | Demographics/Context | Indicator for presence of any invasive line during encounter (binary). |
| hr_max | Vital Signs | Maximum heart rate (beats/min) during observation window. |
| sysbp_min | Vital Signs | Minimum systolic blood pressure (mmHg). |
| diabp_min | Vital Signs | Minimum diastolic blood pressure (mmHg). |
| resp_max | Vital Signs | Maximum respiratory rate (breaths/min). |
| spo2_min | Vital Signs | Minimum peripheral oxygen saturation (%). |
| temp_max_c | Vital Signs | Maximum body temperature ($^{\circ}\text{C}$). |
| temp_delta_6h | Vital Signs | Maximum change in temperature over any 6-hour window ($^{\circ}\text{C}$). |
| max_wbc | Laboratory | Maximum white blood cell count during window. |
| max_neutrophils | Laboratory | Maximum neutrophil percentage. |
| max_anc | Laboratory | Maximum absolute neutrophil count. |
| min_lymphocytes | Laboratory | Minimum lymphocyte percentage. |
| min_hgb | Laboratory | Minimum hemoglobin (g/dL). |
| min_plt | Laboratory | Minimum platelet count ($\times 10^3/\mu\text{L}$). |
| max_glucose | Laboratory | Maximum serum glucose (mg/dL). |
| max_lactate | Laboratory | Maximum serum lactate (mmol/L). |
| max_cr | Laboratory | Maximum creatinine (mg/dL). |
| max_crp | Laboratory | Maximum C-reactive protein (mg/dL). |
| shock_index | Derived | Ratio of maximum heart rate to minimum systolic blood pressure ($\text{shock\_index} = \text{hr\_max} / \text{sysbp\_min}$). |
| fever_flag | Derived | 1 if $\text{temp\_max\_c} \geq 38.0^{\circ}\text{C}$, else 0. |
| hypotension_flag | Derived | 1 if $\text{sysbp\_min} \leq 90$ mmHg, else 0. |
| tachycardia_flag | Derived | 1 if $\text{hr\_max} \geq 100$ beats/min, else 0. |
| tachypnea_flag | Derived | 1 if $\text{resp\_max} \geq 22$ breaths/min, else 0. |
| hypoxia_flag | Derived | 1 if $\text{spo2\_min} \leq 94\%$, else 0. |
| hyperglycemia_flag | Derived | 1 if $\text{max\_glucose} > 127$ mg/dL, else 0. |
| hyperlactate_flag | Derived | 1 if $\text{max\_lactate} > 2.0$ mmol/L, else 0. |
| renal_impairment_flag | Derived | 1 if $\text{max\_cr} > 1.2$ mg/dL, else 0. |
| inflammatory_flag | Derived | 1 if $\text{max\_crp} > 10$ mg/dL, else 0. |
| anemia_flag | Derived | 1 if $\text{min\_hgb} < 12$ g/dL, else 0. |
| thrombocytopenia_flag | Derived | 1 if $\text{min\_plt} < 150 \times 10^3/\mu\text{L}$, else 0. |
| neutrophilia_flag | Derived | 1 if $\text{max\_neutrophils} > 70\%$, else 0. |
| lymphopenia_flag | Derived | 1 if $\text{min\_lymphocytes} < 20\%$, else 0. |
| composite_instability_flag | Derived | Set to 1 if any of fever_flag, hypotension_flag, or hyperlactate_flag are present; otherwise 0. |

**Figure 3.**
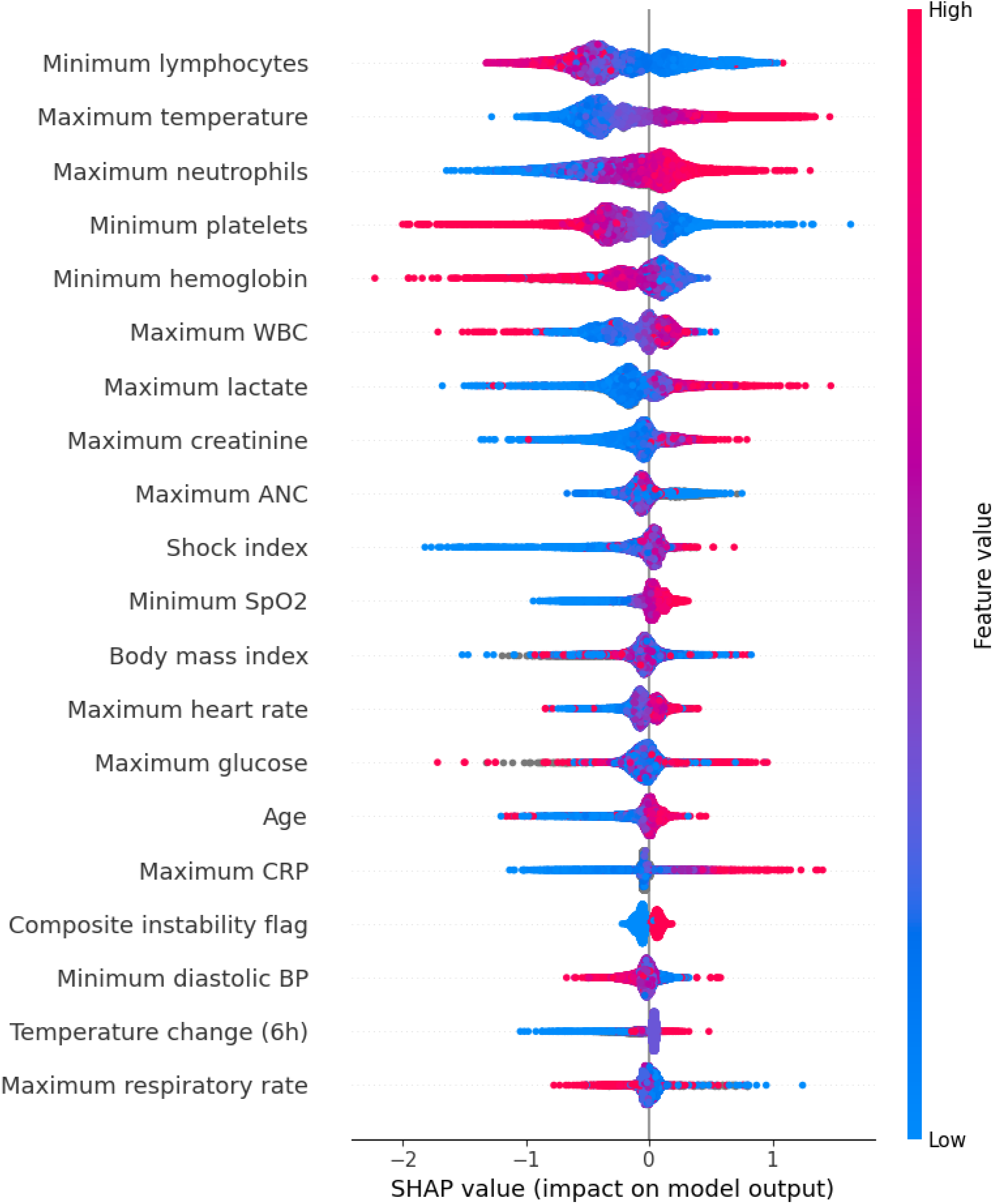
SHAP values showing feature importance for the Cultryx model

## 4 Discussion

The 2024 global shortage of BD BACTEC™ blood culture bottles brought foundational challenges in diagnostic resource allocation to a crisis level, imposing an unprecedented stress test on health-care systems.[4, 5, 6] This reflects the persistent need to balance the imperative to diagnose life-threatening infections against the reality of finite resources. In this study, we demonstrated that the current standards for guiding this balance, whether expert clinical frameworks, their generative AI adaptations, or traditional rule-based heuristics, are insufficient for safe, precise diagnostic stewardship. By leveraging machine learning, we developed and validated Cultryx, a model that outperforms these baselines, offering a mechanism to defer over 26% of blood culture orders while maintaining a sensitivity of 95% for bacteremia. This demonstrates the capability for data-driven methods to deliver better safety and stewardship than existing guidelines and frameworks.

The necessity for a precision-guided approach is highlighted by the blunt impact of the shortage itself. As Lutgring et al. reported, reliance on broad conservation strategies (such as hard stops or single-set limitations) resulted in a 27.4% decrease in culture volume but came at the cost of a 15.3% reduction in the detection of confirmed bloodstream infections.[6] This highlights the critical flaw in non-targeted rationing; without a reliable method to stratify pre-test probability, reducing testing volume invariably risks missing critical diagnoses. Our results suggest that Cultryx bridges this gap, allowing facilities to achieve substantial reductions in volume without the concomitant drop in pathogen detection.

To establish a baseline for current stewardship capabilities, we first benchmarked against the Fabre framework, an expert consensus algorithm that categorizes clinical syndromes based on the reported incidence of bacteremia.[3] While human reviewers successfully applied this framework with high sensitivity (95.7%) in a controlled, retrospective environment (effectively interpreting clinical ambiguity as risk to ensure patient safety) this manual approach is prohibitively labor-intensive for frontline ED workflows, and their specificity remained notably low (35.0%). We attempted to scale this framework using generative AI, but this approach failed. The LLM (GPT-5) achieved a sensitivity of only 71.6% relative to the consensus standard, frequently misclassifying “intermediate risk” patients as “low risk.” This highlights a limitation of current generative AI — while capable of processing complex clinical text, LLMs lack the implicit risk aversion required to safely navigate diagnostic ambiguity in an automated capacity.

Similarly, our evaluation of traditional clinical decision rules confirms their limitations when used as hard constraints for stewardship. The SIRS criteria, designed for broad sepsis screening, lacks the specificity to guide resource rationing, flagging the vast majority of ED encounters as “at risk.” Conversely, the Shapiro Rule proved insufficiently sensitive to serve as a safety net, missing nearly 30% of bacteremic episodes. This suggests that standard clinical rules may not fully capture the variability of bacteremia risk across a heterogeneous ED population. In contrast, Cultryx provided superior overall discrimination (AUROC 0.810). Because it generates a continuous risk score, Cultryx could be explicitly calibrated to a strict 95% sensitivity target, ensuring patient safety while still yielding a 26.2% reduction in culture volume.

The safety profile of Cultryx must be interpreted relative to the actual, rather than an idealized, standard of care. While critics of machine learning interventions often scrutinize any missed bacteremia cases, our analysis demonstrates that established real-world clinical baselines are less safe. As noted above, both the non-targeted rationing of the 2024 shortage and the rigid application of traditional decision aids implicitly accepted reductions in pathogen detection. In contrast, by explicitly calibrating Cultryx to a >95% sensitivity target, this data-driven approach preserves diagnostic integrity while delivering substantial resource savings. Furthermore, safely deferring over 26% of cultures actively mitigates the well-documented downstream harms of diagnostic overuse, which include unnecessary antibiotic exposure, increased hospital length of stay, and adverse events associated with treating contaminants.

To ensure these findings are translatable beyond academic centers with advanced computing infrastructure, we derived Cultryx^score^, a simplified integer-based score. By identifying the highest-yield predictors (e.g., hyperthermia (≥38^°^C), neutrophilia, and thrombocytopenia), we created a transparent bedside tool that retains substantial discriminative power. While Cultryx is operationally superior (saving approximately 3,200 more bottles than Cultryx^score^ at the same safety tier), Cultryx^score^ bridges the digital divide, providing an accessible, empirically validated alternative for immediate implementation in resource-limited settings or during IT downtimes.

Our study has limitations. First, it was conducted at a single academic medical center. While the cohort is large and diverse, external validation is necessary to ensure generalizability to community health settings and to other academic centers. Second, the study is retrospective. While we used a rigorous temporal split to simulate prospective performance, we could not evaluate the clinical outcomes of patients whose cultures would have been deferred, relying instead on high sensitivity targets to infer safety. Third, our outcome labels relied on a rule-based algorithm rather than manual adjudication for the entire dataset. While we implemented strict logic to promote repeated isolates, classification errors remain possible; without granular clinical context, some true pathogens may have been mislabeled as contaminants and vice versa. Fourth, we benchmarked against the Fabre framework to establish an idealized cognitive baseline, which was originally derived for non-neutropenic inpatients. Our application of this framework to an undifferentiated, potentially neutropenic ED population represents an extrapolation. However, we believe this comparison remains valid as the framework’s underlying physiological principles regarding bacteremia risk are broadly applicable. Additionally, our analysis was restricted to encounters where a blood culture was clinically ordered. While this matches the operational goal of diagnostic stewardship (reducing low-value orders), Cultryx’s performance in a broader, undifferentiated patient population not selected for testing remains undefined. Finally, the “bottle savings” estimates assume strict adherence to Cultryx’s recommendations; real-world implementation would require integration into clinical workflows and likely face variable provider compliance.

Although the 2024 global blood culture bottle shortage is over, the crisis exposed a chronic vulnerability in modern medicine: our reliance on high-volume, low-yield testing as a safety net. Returning to pre-crisis baselines effectively means accepting a standard of care where diagnostic overuse and its downstream harms, including unnecessary antibiotics, false positives, and resource strain, are normalized. This study demonstrates that purpose-built machine learning tools like Cultryx offer a precise, scalable alternative to this inefficiency. While expert clinical frameworks provide an idealized baseline, their manual application is operationally prohibitive, and our findings suggest their automation via generative AI currently lacks the necessary safety profile. In contrast, Cultryx provides a viable, automated solution that successfully outperforms established operational clinical rules like SIRS and the Shapiro Rule. By transitioning from crisis-driven rationing to data-driven stewardship, healthcare systems can sustainably elevate their standard of care, reducing low-value testing not just to save bottles, but to improve patient safety and safeguard clinical operations against future supply chain volatility.

## Data Availability

All data produced are available online at Dryad.

https://doi.org/10.5061/dryad.jq2bvq8kp

## Author Contributions

N.P.M. designed the study (including cohort, variables, and prompt engineering), validated data with W.C. and F.A., served as the adjudicating infectious diseases physician for the manual review process, and drafted the manuscript. W.C. and F.A. designed and implemented the methods, developed and executed all study code, performed the statistical analyses, and drafted the manuscript. A.Z. and K.C.B. performed the independent manual chart review of blood culture cases. All remaining authors contributed to the project’s development, reviewed the findings, and provided revisions to the manuscript. All authors approved the final version.

## Use of AI, Chatbots, and Large Language Models

Machine learning (ML) and large language models (LLM) were the subject of this research and were used as described in the manuscript. In addition, commercially available AI-assisted tools were used in a limited way to review and debug code, consistent with common programming practice. These tools were not used to generate novel manuscript text, figures, or results.

## Data Availability

Data available at: doi.org/10.5061/dryad.jq2bvq8kp

## Supplementary Appendix A: Complete List of All Feature Definitions

## Supplementary Appendix B: Full Prompt for Generative AI (GPT-5) Application of the Fabre Framework

### Full GPT-5 Prompt

// Role

You are an infectious diseases physician tasked with classifying an adult patient’s risk of bacteremia at the time of a clinical event or blood culture draw, using criteria from Fabre et al., “Does This Patient Need Blood Cultures? A Scoping Review of Indications for Blood Cultures in Adult Nonneutropenic Inpatients” (DOI: https://doi.org/10.1093/cid/ciaa039).

// Procedure Checklist

Begin with a concise checklist (3-7 bullets) of what you will do; keep items conceptual, not implementation-level.

- Review provided note and EHR data.
- Determine if patient meets exclusion criteria.
- Sequentially check against each risk tier (High, Intermediate, Low) as defined.
- Assign the highest triggered tier or “Undetermined” as appropriate.
- Extract 1-3 verbatim supporting quotes.
- Assess and assign confidence score per guideline.
- Compose and validate concise rationale aligning with Fabre categories/examples.

// Context and Criteria

Scope and Assumptions (strictly enforced):

- Population: Adults (greater than or equal to 18 years), non-neutropenic patients. If data indicate pediatric status or ANC less than 500/µL, return “Undetermined”.
- Use only information present in the note and structured EHR data provided. Do not create or assume data.
- If multiple conditions apply, assign the highest applicable risk tier.

Fabre-Based Risk Tier Definitions (category logic and examples maintained):

- HIGH RISK: Signs of severe sepsis or septic shock, or conditions predisposing to endovascular infection. Examples: catheter-associated bloodstream infection, discitis/native vertebral osteomyelitis, epidural abscess, meningitis, non-traumatic native septic arthritis, ventriculo-arterial shunt infection.
- INTERMEDIATE RISK: Symptoms of systemic infection or localized infections with systemic potential, but without overt severe sepsis. Examples: acute pyelonephritis, cholangitis, non-vascular shunt infections, prosthetic vertebral osteomyelitis, severe community-acquired pneumonia (PSI IV–V). Low-to-intermediate: cellulitis with significant comorbidities, ventilator-associated pneumonia.
- LOW RISK: Non-bacterial syndromes or absence of significant systemic infection signs. Examples: isolated fever/leukocytosis, non-severe cellulitis, lower UTI (cystitis, prostatitis), non-severe community-acquired pneumonia, health-care–associated pneumonia, post-op fever within 48 hrs of surgery.
- Severity interpretation: Use explicit mentions of “severe sepsis,” “septic shock,” vasopressor use, or acute end-organ dysfunction linked to infection. PSI class IV–V corresponds to severe CAP. Do not escalate risk tier without explicit severity cues.

// Decision Algorithm

1. Exclude Out-of-Scope: If pediatric (age ¡18) or neutropenic, classify as “Undetermined”.
2. Check for HIGH RISK: Severe sepsis/septic shock or endovascular-predisposing condition (listed examples or clear equivalents).
3. If not, check INTERMEDIATE RISK: Systemic infection symptoms or localized infection with systemic risk (examples), without severe sepsis.
4. If not, check LOW RISK: Use provided examples and confirm absence of systemic signs.
5. If insufficient or contradictory information, classify as “Undetermined”.
6. If multiple tiers are triggered, assign the highest tier.

After classification, double-check that (a) the highest applicable tier is selected, (b) verbatim quotes support the assigned tier, and (c) rationale accurately references Fabre categories/examples.

// Output Format (strict JSON only)

“Classification”: “HIGH” — “INTERMEDIATE” — “LOW” — “Undetermined”, “Confidence”: number, “Verbatim”: [“quote1”, “quote2”, “quote3”], “Rationale”: “2–4 sentences explaining how the evidence matches the tier, referencing Fabre categories/examples.”

// Output Constraints

- Output strictly as valid JSON. No preamble, markdown, or extra keys.
- Verbatim quotes must be exact excerpts from the input text (180 characters each).
- Confidence scoring: 0.9–1.0 (explicit qualifiers and clear example matches), 0.6–0.8 (probable with partial data), 0.3–0.5 (ambiguous, weak cues), 0.0–0.2 (insufficient or contradicts—likely “Undetermined”).

// Inputs Provided

- Unstructured note text (emergency department provider note).
- Optional structured EHR data (vitals, labs, problem list, etc.).

// Edge Cases Quality Checks

Quality Control Before Submission:

- Ensure classification matches the highest triggered tier.
- Ensure verbatim quotes directly support the assigned tier.
- Ensure rationale is concise and aligns with Fabre categories/examples.
- Output only valid JSON as described above.

